# Spontaneous cerebrovascular reactivity at rest in older adults with and without mild cognitive impairment and memory deficits

**DOI:** 10.1101/2024.06.18.24309109

**Authors:** Allison C Engstrom, John Paul Alitin, Arunima Kapoor, Shubir Dutt, Trevor Lohman, Isabel J Sible, Anisa J Marshall, Fatemah Shenasa, Aimée Gaubert, Farrah Ferrer, Amy Nguyen, David Robert Bradford, Kathleen Rodgers, Lorena Sordo, Elizabeth Head, Xingfeng Shao, Danny JJ Wang, Daniel A Nation

## Abstract

**Background:** Older adults with mild cognitive impairment (MCI) exhibit deficits in cerebrovascular reactivity (CVR), suggesting CVR is a biomarker for vascular contributions to MCI. This study examined if spontaneous CVR is associated with MCI and memory impairment.

**Methods:** 161 older adults free of dementia or major neurological/psychiatric disorders were recruited. Participants underwent clinical interviews, cognitive testing, venipuncture for Alzheimer’s biomarkers, and brain MRI. Spontaneous CVR was quantified during 5 minutes of rest.

**Results:** Whole brain CVR was negatively associated with age, but not MCI. Lower CVR in the parahippocampal gyrus (PHG) was found in participants with MCI and was linked to worse memory performance on memory tests. Results remained significant after adjusting for Alzheimer’s biomarkers and vascular risk factors.

**Conclusion:** Spontaneous CVR deficits in the PHG are observed in older adults with MCI and memory impairment, indicating medial temporal microvascular dysfunction’s role in cognitive decline.

## 1 Introduction

It is increasingly recognized that vascular contributions to cognitive impairment and dementia (VCID) can be prodigious and are often present both in those with and without concomitant neurodegenerative changes such as Alzheimer’s disease^1–5^. Continued efforts to develop and investigate structural imaging markers of large and small vessel diseases relevant to VCID have yielded new insights and tools for earlier detection^6,7^. Deficits in cerebral blood flow (CBF), blood-brain barrier (BBB) permeability and CVR have also been reported in older adults ^8–10^, and patients with MCI exhibit further CBF deficits that include the posterior cingulate and precuneus ^11,12^, increased BBB permeability in the parahippocampal gyrus and hippocampus ^10,13–15^, and deficits in whole brain CVR to hypercapnia^16^.

The study of CVR is of particular interest as a potential marker of VCID given it can be reliably studied non-invasively using MRI and is specific to cerebrovascular function^17,18^. CVR responses can be studied as a measure of cerebrovascular function by inducing hypercapnia through gas inhalation or controlled breathing while simultaneously monitoring end-tidal CO_2_ (_ET_CO_2_), and is commonly quantified as a percent change in CBF per unit change in _ET_CO_2_. In contrast to changes in resting CBF that may reflect either changes in cerebrovascular function or neuronal metabolism or both, CVR responses to blood and CSF CO_2_ levels are thought to be specific to cerebrovascular function^17^. As such, the study of vasodilatory CVR responses is of particular interest as an imaging marker of VCID that may capture early changes in microvascular function that may predate irreversible neuronal injury and cognitive impairment^8,17^.

Fewer studies have examined regional changes in CVR and their relationship to relevant cognitive domains such as memory. One of the few longitudinal studies of CVR reported a decline in CVR in older adults that is related to decline in cognitive functioning and memory, and that CVR decline occurs most rapidly in the temporal lobe^19^. This is consistent with studies showing hippocampal vascularization patterns are related to memory ability in older adults^20^ and that the medial temporal lobe (MTL) may be selectively vulnerable to hypoperfusion^21,22^ Microvascular dysfunction has also been related to early-stage cognitive impairment, independent of traditional vascular risk factors and Alzheimer’s disease biomarker changes^10,13–15^.

Although the majority of research to date has studied CVR responses to experimentally induced hypercapnia, there are also spontaneous fluctuations in CBF at rest that are influenced in part by spontaneous fluctuations in blood and CSF CO_2_ levels^23,24^. These spontaneous fluctuations at rest can be captured by combining either pseudocontinuous arterial spin-labeling (pCASL)- or blood oxygen level dependent (BOLD) signal changes together with capnographic monitoring and are diminished in older adults^9,25^. If spontaneous CVR responses capture microvascular changes contributing to cognitive decline, the relative simplicity and tolerability of this resting state approach would be of great value relative to administration of gas or guided breathing. To our knowledge, no studies to date have examined spontaneous CVR in a well characterized sample of older adults with and without MCI. Based on prior studies of CVR responses to experimentally induced hypercapnia, we hypothesized that spontaneous CVR in global and medial temporal regions would be diminished with age and in older adults with MCI and memory deficits, independent of Alzheimer’s disease biomarker levels.

## 2 Methods

### 2.1 Participants

A total of 161 older adult participants were recruited from the Los Angeles County and Orange County community and all procedures were conducted as part of the Vascular Senescence and Cognition (VaSC) Study at University of Southern California (USC) and University of California Irvine (UCI). Participants were recruited from the community through outreach events, mailing lists, word-of-mouth, online portals, and other modes of community outreach facilitated by the USC Leonard Davis School of Gerontology, and the UCI Alzheimer’s Disease Research Center. All participants were independently living at the time of recruitment, and were aged 55 to 89 years. Study exclusion criteria were a prior diagnosis of dementia, history of clinical stroke, family history of dominantly inherited neurodegenerative disorders, current neurological or major psychiatric disorders that may impact cognitive function, history of moderate-to-severe traumatic brain injury, current use of medications that may impair the central nervous system, current organ failure or other uncontrolled systemic illness, and contraindications for brain MRI. Eligibility for the study was verified via clinical interview and review of current medications with both the participant, and an informant study partner when available.

### 2.2 Standard Protocol Approvals, Registrations, and Patient Consents

The VaSC Study was approved by the USC (HS-14-00784) and UCI (HS-2019-5324) Institutional Review Boards, and all participants gave informed consent prior to participating in the study.

### 2.3 Measures

#### 2.3.1 Neuropsychological Testing and MCI Diagnosis

All participants underwent a clinical interview and comprehensive neuropsychological assessment by a trained technician or doctoral student under the supervision of a licensed clinical neuropsychologist (DAN). The assessment included multiple tests of global cognition, memory, attention/executive function, and language. The Mattis Dementia Rating Scale was also used to screen participants for dementia^26^. Participants were classified as cognitively unimpaired (CU) if they did not meet published neuropsychological criteria for MCI^27^. Specifically, MCI was diagnosed when participants scored >1 standard deviation below demographically-referenced z-scores on >1 measure within a domain or >2 measures across domains. The neuropsychological battery used for diagnosis utilized 3 tests per domain of function. Participants with impairment on >1 memory measure were considered amnestic MCI. Numerous studies have previously established the validity of this neuropsychological approach to MCI^28–30^. All neuropsychological testing and diagnostic assessment were conducted blinded to all clinical, biomarker and imaging findings.

In addition, a memory composite was created by averaging the demographically corrected z-scores from the memory tests which included: Logical Memory 2 (LM2), Rey Auditory Verbal Learning Test Trial 7 (RAVLT7), and Rey Auditory Verbal Learning Test Recognition (RAVLT Recognition).

#### 2.3.2 Neuroimaging and Cerebral Perfusion

All participants underwent a brain MRI on a 3T scanner (Siemens MAGNETOM Prisma System). The following sequences were examined for the current analysis: 3D T1-weighted anatomical scan for qualitative assessment of brain structures and abnormalities (Scan parameters: TR = 2300 ms; TE = 2.98 ms; TI = 900 ms; flip angle = 9 deg; FOV = 256 mm; resolution = 1.0 × 1.0 × 1.2 mm3; Scan time = 9 minutes), and 3D gradient and spin-echo (GRASE) pCASL for CBF quantification. The scan parameters were as follows for pCASL: TR = 5000 ms; TE at University of Southern California = 36.3 ms; TE at University of California, Irvine = 37.46 ms; FOV = 240 mm; resolution = 2.5 × 2.5 × 3.4 mm3; slice thickness = 3.42 mm; number of slices = 24; labeling duration = 1517 ms; post-labeling delay = 2000 ms. There was a total of 32 acquisitions (1 M0 image + 1 dummy image + 30 alternating tag and control images), with a total scan time of 5 minutes 25 seconds, yielding 15 tag-control pair images.

The pCASL scans were pre-processed using the ASLtbx pipeline, implemented in SPM12 within MATLAB^31,32^. Pre-processing steps for pCASL scans included motion correction, co-registration to individual participants’ structural T1-weighted image, spatial smoothing with a 6 mm full-width at half-maximum Gaussian kernel, and tag-control subtraction resulting in 15 tag-control pairs for each participant with values for absolute CBF (mL/100g tissue/min). All CBF images were thresholded below 10 or above 150mL/100g/min to exclude CBF outside the expected physiological range of gray matter ^33,34^. Tag-control pairs were warped to MNI space and averaged to create mean CBF maps for each participant. Resulting mean CBF maps were visually inspected for quality and gross abnormalities (i.e., large signal dropout). Partial volume correction was performed by applying participant-specific gray matter masks derived from the gray matter tissue class segmentation of T1-weighted structural images^35^. Segmented gray matter maps were thresholded at 0.3, binarized, and multiplied by the mean CBF maps to ensure CBF was limited to gray matter.

Capnography indexed _ET_CO_2_ during MRI acquisition using an M3015A sidestream CO_2_ extension module (Philips Medical Systems) connected to a nasal cannula into which participants breathed. To correct for sampling tubing latency, _ET_CO_2_ time series were shifted by a pre-calibrated duration of time (i.e., 10 seconds for the present study). For the baseline pCASL scan, _ET_CO_2_ was extracted from the raw time series in accordance with the breathing rate (i.e., at every expiration). Participants were excluded from analysis if they failed to adhere to breathing instructions (e.g., lack of positive peaks in raw data – participant might be breathing through the mouth).

#### 2.3.4 CVR Maps

Spontaneous CVR has been quantified variously in the literature^23^. For consistency with prior studies utilizing our paradigm, CVR was conceptualized as the percent change in CBF per unit change in _ET_CO_2_. Specific computational approach has been previously outlined. In the present study, participant’s individual CVR maps were generated using resting state CBF with simultaneous capnography with the following equation (1) computed in each voxel:

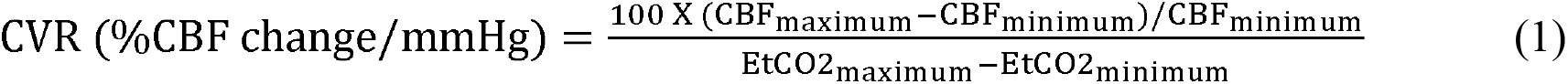

Based on prior studies^13,16^ and our hypothesized relations with MCI and memory impairment, we examined spontaneous CVR in the whole brain and in two medial temporal structures (hippocampus and PHG) (Figure 1).

**Figure 1:**
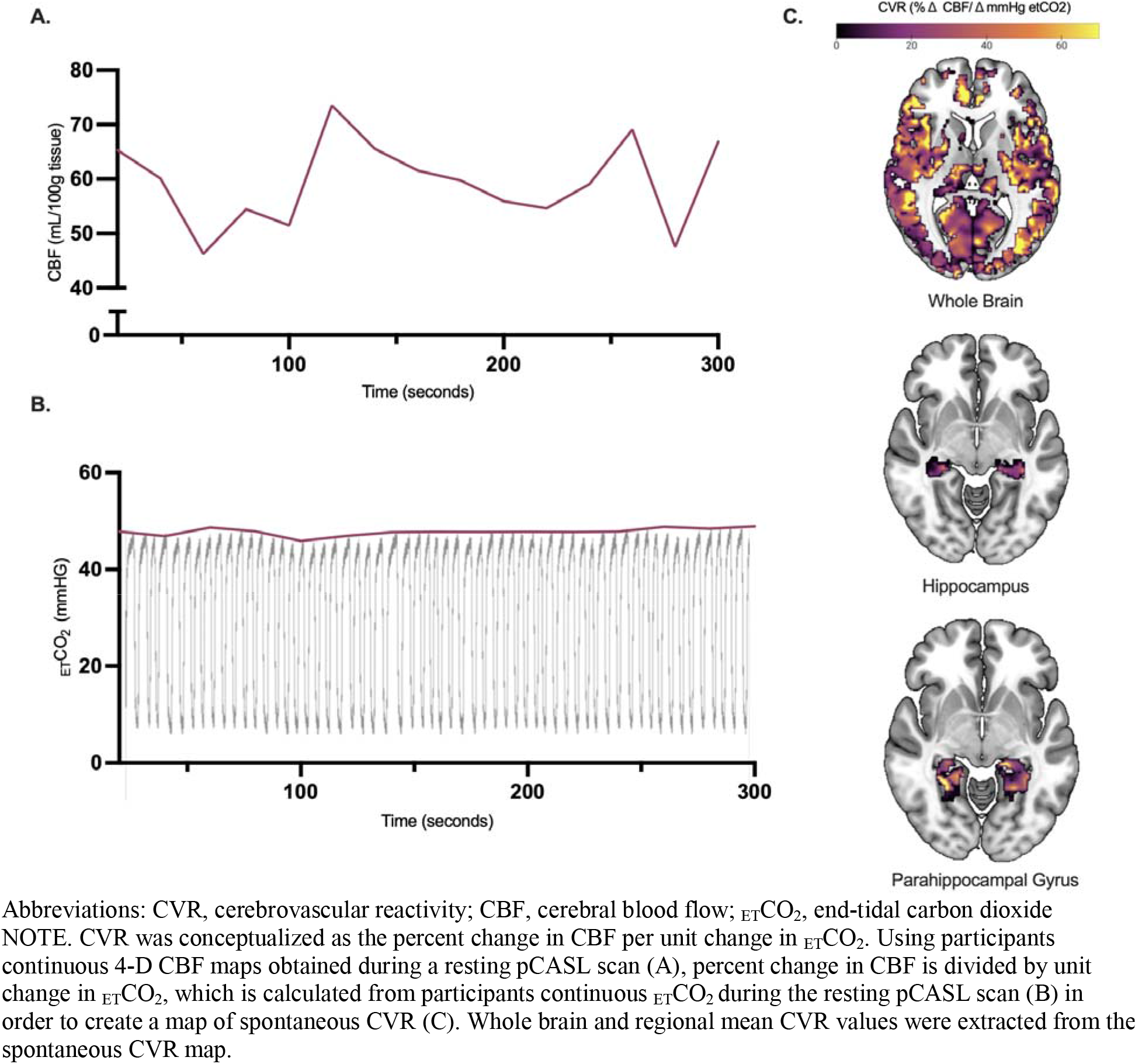
Spontaneous CVR mapping methodology.

#### 2.3.5 Vascular Risk Factors (VRFs)

The participants’ VRF burden was evaluated through clinical interviews with the participant and a knowledgeable informant (when available) and included a history of cardiovascular disease (heart failure, angina, stent placement, coronary artery bypass graft, intermittent claudication), hypertension, hyperlipidemia, type 2 diabetes, smoking, atrial fibrillation, and transient ischemic attack or minor stroke. The total VRF burden was defined by the sum of these risk factors and then dichotomized into lower vs. higher burden (0-1 vs. 2+ VRFs) based on prior studies^13,39,40^.

#### 2.3.6 Plasma AD Biomarkers

Participants underwent venipuncture after an overnight fast, and blood plasma was separated by centrifugation and stored at -80°C until AD biomarker assays. Plasma phosphorylated tau (pTau_181_) and amyloid beta (A _40_ and A _42_) concentrations were obtained using the digital immunoassay, single molecule array (Simoa) pTau_181_ Advantage v2.1 and Neurology 3-Plex A (N3PA) Advantage Kits, respectively, conducted in the same lab (EH).

#### 2.3.7 *APOE* Genotyping

Blood samples were used to determine participant *APOE* genotype, as previously described_41_. Briefly, genomic DNA was extracted using the PureLink Genomic DNA Mini Kit (Thermo). The isolated DNA concentration was determined using a NanoDrop One (Thermo). DNA was then stored at_−_80 °C for long-term storage. Isolated DNA was first diluted to a concentration of 10 mg/μL. PCR reactions were performed in a final volume of 25 μL containing 25 ng DNA, 0.5 μM of both forward and reverse primers (forward: ACGGCTGTCCAAGGAGCTG; reverse: CCCCGGCCTGGTACACTG), and 1×L Master Mix (Qiagen) diluted in H2O. For the amplification, a T100 Thermal Cycler (BioRad) was used with the following settings: 95 °C for 10 min; 32 cycles of 94 °C for 20 s, 64 °C for 20 s, and 72 °C for 40 s; followed by 72 °C for 3 min. 15 μL of the DNA PCR product was digested with Hhal-fast enzyme at 37 °C for 15 min. The digested PRC product was added to a 3% agarose gel in 1×_borax buffer for gel electrophoresis. The gel was run at 175 V for 25 min and visualized on ChemiDoc (BioRad) with a GelRed 10,000×_gel dye. *APOE4* carrier status was defined as *APOE4* carriers (at least one copy of the ∑4 allele) or *APOE4* non-carriers (no copies of the ∑4 allele).

### 2.4 Data Availability

The data that support the findings of this study are available upon reasonable request from the corresponding author, DAN, based on the conditions outlined by the Data Availability Policy and Statement.

### 2.5 Data Analysis

A total of 161 participants were studied and characterized by demographics, spontaneous whole brain CVR and MCI data for statistical analysis (Table 1). Spontaneous CVR in the hippocampus and PHG was available on a subset of participants with usable data after exclusion of negative values and outlier removal (N = 150). Spontaneous CVR values (whole brain, hippocampus, and PHG) were normally distributed. Outliers of whole brain, hippocampus and PHG spontaneous CVR were screened and removed if they were greater than +/-3 SD from the mean, with a total of 2 outliers removed.

**Table 1.** Participant demographics.

| **Table 1.** Participant demographics |  |
| --- | --- |
| ***N* = 161** | **M (SD) or n(%)** |
| **Age** | 69.5 (7.6) |
| **Sex, Female, n (%)** | 105 (65.2) |
| **Education** | 16.5 (2.4) |
| **Race, n (%)** |  |
| White | 127 (78.9) |
| Black or African American | 4 (2.5) |
| American Indian or Alaska Native | 2 (1.2) |
| Native Hawaiian or other Pacific Islander | 1 (0.6) |
| Asian | 23 (14.3) |
| Other | 4 (2.5) |
| **Ethnicity** |  |
| Hispanic or Latino | 8 (5.0) |
| Non-Hispanic or Latino | 153 (95.0) |
| **Cognitive Impairment, n (%)** |  |
| MCI | 30 (18.6) |
| Amnesic MCI | 16 (9.9) |
| Cognitively Unimpaired | 131 (81.4) |
| <sup>1</sup>***APOE4* Carriers (n, 3/4, 4/4 %)** | 66 (47.1) |
| <sup>2</sup>**p-Tau<sub>181</sub>** | 24.1 (12.0) |
| <sup>3</sup>**Aβ<sub>42/40</sub> ratio** | 0.04 (0.02) |
| **Vascular risk factors, n (%)** |  |
| Hypertension | 61 (37.9) |
| Hyperlipidemia | 74 (46.3) |
| Diabetes | 20 (12.4) |
| Smoking | 56 (34.8) |
| Cardiovascular Disease | 17 (10.6) |
| Atrial Fibrillation | 8 (5.0) |
| Transient Ischemic Attack | 3 (1.9) |

Abbreviations: Aβ , amyloid beta; MCI, mild cognitive impairment; p-Tau, phosphorylated tau
<sup>1</sup>Subset of n = 140 Participants, <sup>2</sup>Subset of n = 114 Participants, <sup>3</sup>Subset of n = 111 Participants

Linear regression analyses were conducted, with age as the predictor and spontaneous CVR as the outcome variable, to determine the relationship between spontaneous CVR and age, for both whole brain hippocampus, and PHG spontaneous CVR. The relationship between CBF and age was also evaluated to determine if spontaneous CVR was indicative of microvascular change above what is evident in CBF. All neuropsychological test scores used in the diagnosis of MCI were demographically corrected for age, sex and education, therefore, independent samples *t*-tests were performed to determine the relationship between hippocampus and PHG spontaneous CVR in global cognitive impairment. An additional, independent means *t*-test was conducted to evaluate the relationship between hippocampus and PHG spontaneous CVR and memory impairment by comparing hippocampal and PHG CVR in those with amnestic MCI and participants without cognitive impairment.

The relationship between spontaneous CVR in the hippocampus and the PHG was compared in participants with and without amnesic MCI (defined by impairment on at least two memory tests). Linear regression was performed with hippocampal and PHG spontaneous CVR and a memory composite. In addition, an independent means *t*-test was performed to compare hippocampal and PHG spontaneous CVR in participants with memory composite z-scores of < -1 to those with composite z-scores of > -1. Lastly, in order to determine the relationship between individual memory tests (LM2, RAVLT7 and RAVLT Recognition) and hippocampal and PHG spontaneous CVR, linear regression was conducted using raw scores and controlling for age, sex and education.

Lastly to account for multiple comparisons, False Discovery Rate (FDR) correction was performed^38^.

## 3 Results

Linear regression revealed a significant negative association between age and whole brain spontaneous CVR [B = -0.41, 95% CI (-0.65, -0.17), *p* = .00096], and this relationship was not attenuated by adjustment for sex. In a regression model with age and whole brain CBF, there was no significant association [B *=* 0.09 95% CI (-.05, 0.23), *p* = .646]. PHG spontaneous CVR was also not significantly associated with age [B = -0.142, 95% CI (-0.52, .03), *p* = .083], and PHG CBF did not show a significant association with age [B = 0.109 95% CI (-.05, 0.23), *p* = .186] (Figure 2). There was no significant relationship between hippocampal spontaneous CVR and age.

**Figure 2:**
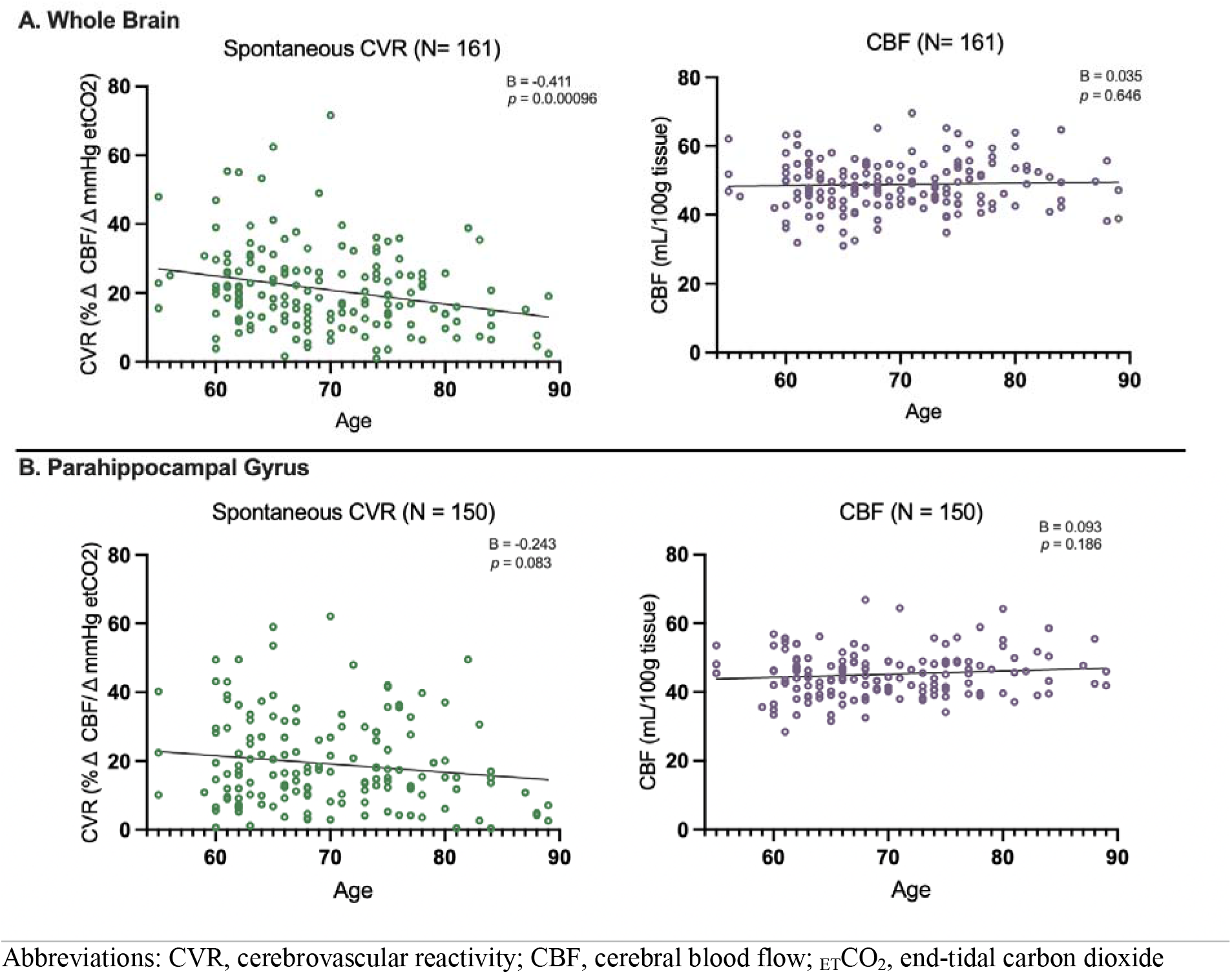
Regression plots of the relationship between age and spontaneous cerebrovascular reactivity (CVR) compared to that of age and cerebral blood flow (CBF) for A: whole brain, and B: parahippocampal gyrus (PHG).

Spontaneous PHG CVR was significantly lower in participants with MCI (M = 14.27, SD = 9.28) than those who were cognitively unimpaired (M = 20.24, SD = 13.87) [*t* (61.8) *=* 2.90 95% CI (1.93, 10.46), *p* = .005], but there was no difference in PHG CBF between MCI and cognitively unimpaired (Figure 3 A and B). Spontaneous PHG CVR was significantly lower in participants with amnestic MCI (M = 12.68 SD = 9.08) compared to those who were cognitively unimpaired (M = 20.62 SD = 13.86), [*t*(25.46) = 3.055 95% CI (2.60, 13.29), *p* = .005] (Figure 3 C). There was no significant relationship between hippocampal spontaneous CVR and MCI or amnestic MCI.

**Figure 3:**
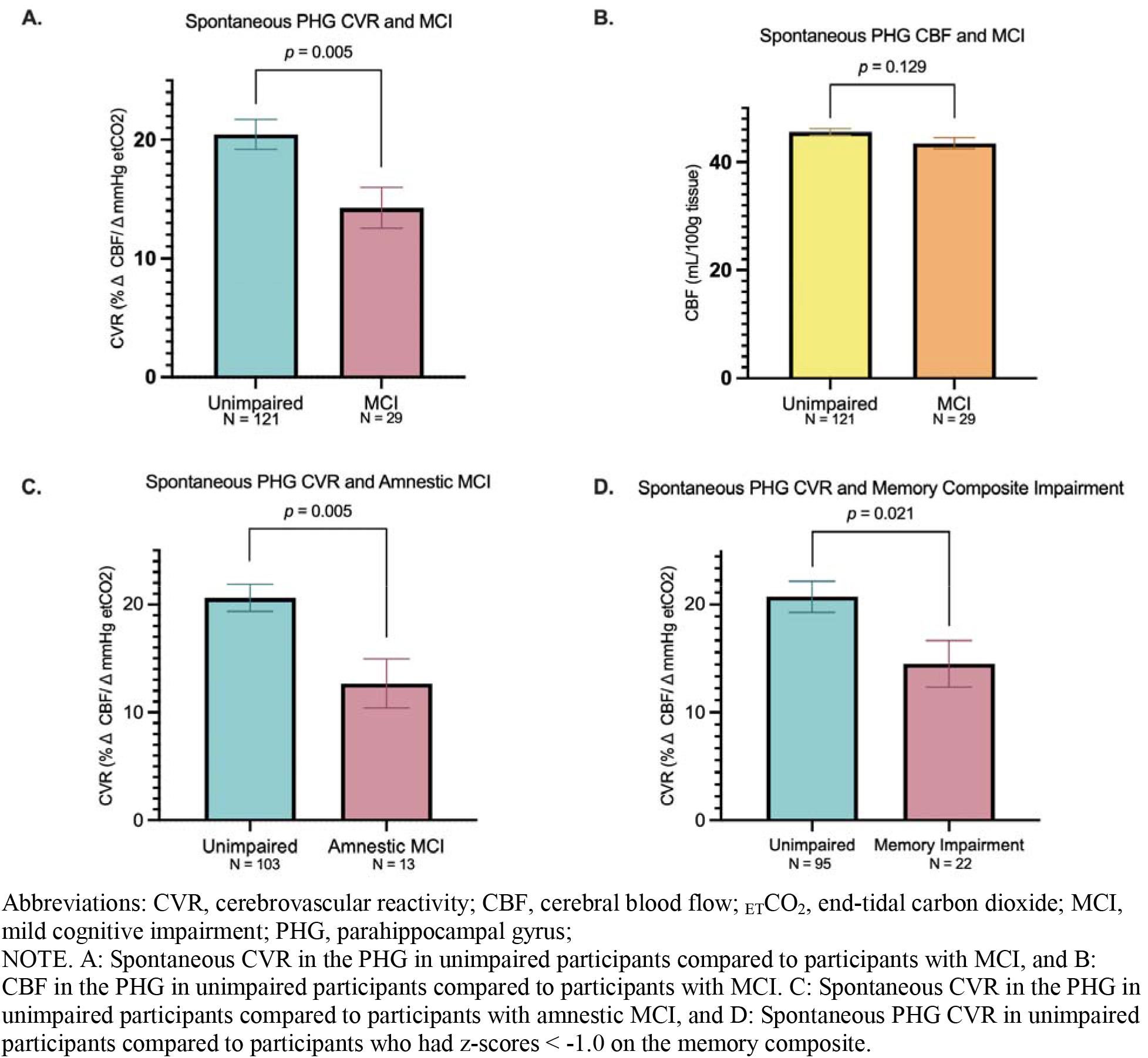
Spontaneous PHG CVR and Cognitive Impairment.

Spontaneous PHG CVR was significantly lower in participants with impaired (z-score < - 1) memory composite z-scores (M = 14.50 SD = 10.09) compared to those with memory composite z-scores > -1 (M = 20.72 SD = 14.13) [*t*(42.42) = 2.40 95% CI (0.98, 11.45), *p* = .021] (Figure 3D). In linear regression, spontaneous PHG CVR was positively associated with memory composite z-scores [B = 0.255, 95% CI (0.56, 3.12), *p* =.005] (Table 2). There was no significant relationship between hippocampal spontaneous CVR and memory composite impairment.

**Table 2:** Break down of regression analysis results investigating the relationship between spontaneous PHG CVR and neuropsychological test of memory including: a memory composite, RAVLT 7, RAVLT Recognition, and Logical Memory 2

| Model | R | $\Delta R^2$ | *df* | $\beta$ | *p* value |
| --- | --- | --- | --- | --- | --- |
| PHG CVR and Memory Composite | 0.255 | 0.065 | (1,116) | 0.255 | 0.005* |
| PHG CVR and RAVLT trial 7 | 0.301 | 0.090 | (4,113) |  | 0.029* |
| RAVLT 7 raw score |  |  |  | 0.277 | 0.007* |
| Age |  |  |  | -0.046 | 0.643 |
| Sex |  |  |  | 0.069 | 0.467 |
| Education |  |  |  | 0.082 | 0.403 |
| PHG CVR and RAVLT Recognition | 0.226 | 0.051 | (4, 134) |  | 0.133 |
| RAVLT recog raw score |  |  |  | 0.182 | 0.046* |
| Age |  |  |  | -0.115 | 0.187 |
| Sex |  |  |  | 0.019 | 0.830 |
| Education |  |  |  | 0.062 | 0.498 |
| PHG CVR and LM2 | 0.248 | 0.062 | (4, 113) |  | 0.124 |
| LM2 raw score |  |  |  | 0.188 | 0.055 |
| Age |  |  |  | -0.110 | 0.256 |
| Sex |  |  |  | 0.010 | 0.915 |
| Education |  |  |  | 0.092 | 0.360 |

Abbreviations: CVR, cerebrovascular reactivity; LM2, Logical Memory 2; PHG, parahippocampal gyrus; RAVLT, Rey Auditory Verbal Learning Test

Linear regression evaluating the relationship between spontaneous PHG CVR and raw scores on memory tests revealed a positive association between participant performance on delayed story recall (LM2) and spontaneous PHG CVR [B = 0.188, 95% CI (-0.01, 0.63), *p* = .055]. There was also a significant positive association between participant performance on delayed word list recall (RAVLT trial 7) and spontaneous PHG CVR [B = 0.277, 95% CI (0.27, 1.69), *p* = .007]. Lastly there was also a significant positive association between participant performance on delayed word list recognition (RAVLT Recognition) and spontaneous PHG CVR [B = 0.182, 95% CI (0.02, 2.03), *p* = .046]. All analyses were controlled for age, sex and education (Table 2).

All primary analyses were included for FDR correction. All significant findings survived FDR correction except for the relationship between PHG spontaneous CVR and raw scores on RAVLT recognition.

### 3.1 Sensitivity Analysis

Consistent results were observed in models controlling for 1) plasma Aβ and 2) vascular risk factors, and in models controlling for 1) p-Tau_181_ and 2) vascular risk factors, with the exception of the relationship between PHG spontaneous CVR and memory composite impairment (Supplementary Tables 1 and 2). In models with amyloid beta and vascular risk factors the relationship between PHG spontaneous CVR and memory composite impairment was attenuated to *p* = .08.

## 4 Discussion

The present study found that deficits in spontaneous PHG CVR at rest are associated with cognitive impairment, and in particular with amnestic cognitive impairment and deficits on multiple tests of delayed verbal memory. These findings are consistent with previously reported deficits in experimentally-induced CVR related to cognitive impairment and memory decline, and further extend these findings to spontaneous CVR at rest^16,19,42,43^. Prior studies have noted microvascular dysfunction in the hippocampus and PHG in older adults with cognitive impairment, including vulnerability to hypoperfusion and blood brain barrier breakdown^13,44^, but few CVR studies have examined medial temporal regions related to memory impairment. Importantly, the study findings were largely unchanged after controlling for traditional vascular risk factors and plasma AD biomarkers, and were not accounted for by differences in resting CBF. Together these findings suggest cerebrovascular dysfunction within medial temporal regions are observed during early-stage cognitive and memory impairment, independent of AD pathophysiologic change. Additional research is needed to determine whether spontaneous CVR at rest may be a valuable early-stage biomarker of vascular contributions to cognitive decline.

Although whole brain spontaneous CVR was correlated with age, there were no whole brain differences between MCI and cognitively unimpaired participants. Conversely, PHG spontaneous CVR was not significantly correlated with age, but was decreased in MCI and memory impairment. Prior studies using experimentally-induced CVR approaches have noted age effects both on whole brain and temporal lobe CVR to hypercapnia^8,16^. Further studies are needed, but the findings of the present study could suggest that deficits in spontaneous PHG CVR are indicative of pathological versus normal aging.

The potential value of spontaneous PHG CVR as a biomarker is in part due to its relative simplicity and tolerability when compared with other methods requiring manipulation of blood and CSF CO_2_ levels. Prior studies have evaluated spontaneous CVR using various methods, often focused on resting state BOLD fluctuations^23,45–47^. However, the ability of spontaneous CVR to capture adequate changes in _ET_CO_2_ at rest has been questioned^48–50^. Our approach focused on resting state pCASL which has the advantage of being a straightforward measure of CBF, but has the disadvantage of low temporal resolution and signal-to-noise ratio. Nevertheless, the present study found that calculation of CVR with spontaneous CBF and _ET_CO_2_ changes at rest using standard methods did in fact provide useful information beyond CBF alone. Additional studies using other modalities, including BOLD, and quantification methods will provide further insights into the relative advantages of different approaches to spontaneous regional CVR at rest.

Some limitations of this study include the cross-sectional design and relative lack of diversity in the sample, which may limit the generalizability of the results. The present study also only evaluated CBF and CVR in the grey matter only as the current sequence was not optimized to capture white matter CBF. Moreover, in this study we were able to assess spontaneous CVR using pCASL. Given that participants in the study had no history of stroke, dementia, or other neurological condition, additionally studies are needed to determine if deficits in spontaneous CVR are associated with more severe cerebrovascular and neurodegenerative conditions. Longitudinal studies are also warranted to examine the predictive value of spontaneous CVR as a preclinical biomarker of cognitive decline.

## Data Availability

All data produced in the present study are available upon reasonable request to the authors

## Acknowledgements

This research was supported by National Institutes of Health grants (DAN: R01AG064228, R01AG060049, R01AG082073, P01AG052350, P30AG066530), (EH: UCI ADRC P30AG066519) and the Canadian Institutes of Health Research (AK: DFD-170763)

## Non-standard abbreviations

BBB: blood-brain barrier
CBF: cerebral blood flow
CVR: cerebrovascular reactivity
_ET_CO2: end-tidal carbon dioxide
LM2: Logical Memory 2
MCI: mild cognitive impairment
MTL: medial temporal lobe
PHG: parahippocampal gyrus
pCASL: pseudo-continuous arterial spin labeling
RAVLT: Rey Auditory Verbal Learning Test
VRFs: vascular risk factors

## 5 Acknowledgments, Sources of Funding, and Disclosures

This research was supported by National Institutes of Health grants (DAN: R01AG064228, R01AG060049, R01AG082073, P01AG052350, P30AG066530), (EH: UCI ADRC P30AG066519) and the Canadian Institutes of Health Research (AK: DFD-170763). Authors had no conflicts of interest.

