## Supplemental Tables 1 and 2 for "Spontaneous cerebrovascular reactivity at rest in older adults with and without mild cognitive impairment and memory deficits"

Supplementary Tables

Supplementary Table 1

Linear regression model estimates with additional covariates.

| Added covariates | | **ß (95% CI)** |
| --- | --- | --- |
| Whole Brain Spontaneous CVR & Age | | |
| **Aβ_42/40_ and Vascular Risk Factors** | -0.27 (-0.68, -.013) | |
| **p-Tau_181_ and Vascular Risk Factors** | -0.78 (-0.70, -.012) | |
|  | PHG Spontaneous CVR & Age | |
| **Aβ_42/40_ and Vascular Risk Factors** | -.020 (-0.58, 0.00) | |
| **p-Tau_181_ and Vascular Risk Factors** | -.020 (-0.60, 0.01) | |
|  | PHG Spontaneous CVR & Memory Composite | |
| **Aβ_42/40_ and Vascular Risk Factors** | 0.25 (0.17, 2.90) | |
| **p-Tau_181_ and Vascular Risk Factors** | 0.22 (0.04, 2.70) | |

VRF = vascular risk factors

Supplementary Table 2

Analysis of covariance (ANCOVA) model estimates with additional covariates.

| Added Covariates | *F* | *df* | 95% CI | *p* | η_p_^2^ |
| --- | --- | --- | --- | --- | --- |
|  | PHG Spontaneous CVR & MCI | | | | |
| **Aβ_42/40_ and Vascular Risk Factors** | 4.23 | 1, 96 | 0.23, 11.30 | 0.04 | 0.04 |
| **p-Tau_181_ and Vascular Risk Factors** | 3.84 | 1, 99 | -0.07, 11.05 | 0.05 | 0.04 |
|  | PHG Spontaneous CVR & Amnestic MCI | | | | |
| **Aβ_42/40_ and Vascular Risk Factors** | 3.98 | 1, 86 | 0.02, 14.60 | 0.05 | 0.04 |
| **p-Tau_181_ and Vascular Risk Factors** | 3.94 | 1, 89 | 0.00, 14.34 | 0.05 | 0.04 |
|  | PHG Spontaneous CVR & Memory Composite Impairment | | | | |
| **Aβ_42/40_ and Vascular Risk Factors** | 3.15 | 1, 75 | -0.75, 12.89 | 0.08 | 0.04 |
| **p-Tau_181_ and Vascular Risk Factors** | 2.55 | 1, 78 | -0.27, 0.18 | 0.08 | 0.04 |

VRF = vascular risk factors
